# Supporting COVID-19 Policy-Making with a Predictive Epidemiological Multi-Model Warning System

**DOI:** 10.1101/2020.10.18.20214767

**Authors:** Martin Bicher, Martin Zuba, Lukas Rainer, Florian Bachner, Claire Rippinger, Herwig Ostermann, Nikolas Popper, Stefan Thurner, Peter Klimek

## Abstract

In response to the SARS-CoV-2 pandemic, the Austrian governmental crisis unit commissioned a forecast consortium with regularly projections of case numbers and demand for hospital beds. The goal was to assess how likely Austrian ICUs would become overburdened with COVID-19 patients in the upcoming weeks. We consolidated the output of three independent epidemiological models (ranging from agent-based micro simulation to parsimonious compartmental models) and published weekly short-term forecasts for the number of confirmed cases as well as estimates and upper bounds for the required hospital beds. Here, we report on three key contributions by which our forecasting and reporting system has helped shaping Austria’s policy to navigate the crisis, namely (i) when and where case numbers and bed occupancy are expected to peak during multiple waves, (ii) whether to ease or strengthen non-pharmaceutical intervention in response to changing incidences, and (iii) how to provide hospital managers guidance to plan health-care capacities. Complex mathematical epidemiological models play an important role in guiding governmental responses during pandemic crises, in particular when they are used as a monitoring system to detect epidemiological change points.

## I. INTRODUCTION

The first known COVID-19 cases in Austria appeared at the end of February 2020 together with one of the first European superspreading events in the Tyrolean tourist region of Ischgl, visited by travellers from all over the globe [1]. In the first half of March 2020, a nation-wide spread of the virus occurred with an exponential rise of confirmed cases [2]. These developments occurred against the dramatic backdrop of the neighboring country of Italy, where despite strict non-pharmaceutical interventions (NPIs) case numbers kept surging, hospital capacities were exceeded and the military had to step in to remove piling bodies [3, 4]. To understand how likely similar developments would have been in Austria, mid-March a forecast consortium was formed and tasked by the government with a weekly forecasting of the expected developments in case numbers and how these developments would translate into demand for healthcare resources. The overarching policy goal at this stage was to navigate the crisis without overburdening the Austrian healthcare system. Over the summer, this was also given a legal basis with a clause in the Austrian COVID law that stay-at-home orders may only be implemented if healthcare capacities are in danger of becoming exhausted [5]. The Austrian Corona Commission, an advisory committee to the minister of health tasked with assessment of epidemiological risk, defined that ICUs would be able to cope with situations in which up to 33% of all ICU beds would be occupied by COVID-19 patients [6].

At an earlier stage than other middle European countries, Austria took a series of non-pharmaceutical interventions (NPIs) in response to the crisis [7] during the first wave. Next to a ramping up of healthcare and public health capacities, airport restrictions and landing bans intensified in the first week of March. Gatherings were limited to 500 persons, cultural and other events started to be cancelled on March 10. On March 16, Austria went into a full lockdown with schools, bars, restaurants, and shops being closed, as well as a transitioning into home office for all non-essential employees [7]. Together with other, earlier measures, these NPIs effectively led to a rapid reduction of daily infection numbers. The number of new cases per day reached a first peak on March 26 with 1,065 cases [8]. In the first wave, COVID-19 related hospitalisations peaked on March 31 with 912 regular beds, whereas the ICU utilization peaked on April 8 with 267 beds being occupied by COVID-19 patients. Daily new cases decreased over April after which they fluctuated at values below one hundred until July [9].

Starting in July 2020 case numbers in Austria started to increase again leading to a second wave in October. In response to this rise of case numbers, the Austrian government implemented a series of lockdowns with varying levels of stringency since beginning of November 2020.Hospitalisations peaked in the end of November, with 3,985 regular beds occupied on November 24, and 709 ICU beds occupied on November 25, respectively. According to officially reported data, this peak brought Austrian hospitals very close to the critical limit of 33% ICU bed occupancy by COVID patients.

The Austrian COVID-19 forecast consortium provided weekly short-term forecasts for case numbers and required hospital beds. In particular, our role was to forecast how likely the 33% threshold of ICU beds being occupied by COVID-19 patients would be crossed within our forecast horizon. Our consortium consisted of three independent modelling teams with experience in the use and development of mathematical and computational models to address epidemiological and public health challenges [10–16]. The consortium was complemented with experts from the Ministry of Health, the Austrian Agency for Health and Food Safety, as well as external public health experts in weekly meetings.

A plethora of epidemiological models to forecast the spread of COVID-19 has been proposed recently [17–22]. In the forecast consortium, we consolidated the output of three models into a single forecast of case numbers for 8–14 days and used these case numbers to predict the numbers of required hospital and ICU beds for 21 days for the country as a whole and for each of its nine federal states. In addition to these point estimates, we also provided upper and lower bounds for these numbers at various levels of uncertainty. These upper bounds of the hospital bed forecasts served as a guidance system for the regional hospital managers, allowing them to estimate how many beds should be reserved for COVID-19 patients if they were willing to accept a given level of risk. These forecasts have been published each week on the homepage of the Ministry of Health. [23].

In this work we present the forecast and reporting system we developed based on the three independent forecasting models. After a brief summary of the individual models and strategies to combine their output, we describe the accuracy of our forecasts and how this accuracy depended on the phase of the epidemics (i.e., in high or low incidence phases, during waves, etc.). We discuss how our results were received by policy-makers, stakeholders in the healthcare system, and the public. We outline the main contributions of our approach to chart a safe path to re-open the country after the first lockdown and how the system informed the necessity for a second lockdown in November 2020. The aim of this work is to communicate the methods applied and developed which allowed three individually thinking modelling and simulation research units to work together in a joint task force producing a consolidated forecast, the benefits and shortcomings of the process, and the political impact of the achieved results. We conclude that epidemiological models can be useful as the basis for short-term forecast-based monitoring systems to detect epidemiological change points which in turn inform on the necessity to strengthen or ease NPIs.

## II. METHODS

We used three conceptually different epidemiological COVID-19 models, developed and operated individually by three research institutions, namely a modified SIR-X differential equation model (Medical University of Vienna / Complexity Science Hub), an Agent-Based simulation model (TU Wien / dwh GmbH), and a simplified state space model (Austrian National Public Health Institute).

### A. Data

Although the three models use different parameters and parametrization routines, they are calibrated using the same data to generate weekly forecasts. Consequently, differences between the model forecasts are a result of different model structure and calibration, but not a result of different data sources. The models also used different nowcasting approaches to correct for late reporting of positive test results. We used data from the official Austrian COVID-19 disease reporting system (EMS, [9]). The system is operated by the Austrian Ministry of Health, the federal administrations, and the Austrian Agency for Health and Food Safety.

For every person tested positively in Austria, the EMS contains information on the date of the test, date of recovery or death, age, sex and place of residence. Furthermore, hospital occupancy of COVID-19 patients in ICU and normal wards are available from daily reports collected by the Ministry for Internal Affairs.

### B. Extended SIR-X Model

One of our models is an extension of the recently introduced SIR-X model [17]. The original SIR-X model introduced a parsimonious way to extend the classic SIR dynamics with the impact of NPIs. In particular, two classes of NPIs are considered. First, there are NPIs that lead to a contact reduction of *all* individuals (susceptible and infected ones). Such NPIs include social distancing and other lockdown measures. Second, the model also represents NPIs that reduce the effective duration of infectiousness for infected individuals. Contact-tracing and quarantine belong to this category.

The original SIR-X model does not offer a way to model the taking back of NPIs. We extended the model by introducing a mechanism by which susceptible but quarantined individuals increase their number of contacts again; a model we dub the XSIR-X model, see SI Appendix A. Further, we structured the population according to age, introduced multiple calibration phases to model behavioural changes in the population over time, and used mobility data to identify such turning points [24]. Forecast errors are estimated by recalibrating the model to perturbed data points that are displaced proportionally to the empirical deviation between model and data; see SI Appendix A.

### C. Agent-Based SEIR Model

The second model is an Agent-Based SEIR type model [13], furthermore abbreviated as AB model. It is stochastic, population-dynamic and depicts every inhabitant of Austria as one model agent. It uses sampling methods to generate an initial agent population with statistically representative demographic properties and makes use of a partially event-based, partially time-step (1 day)-based update strategy to enhance in time.

It is based on a validated population model of Austria including demographic processes like death, birth, and migration [15]. Contacts between agents are responsible for disease transmission and are sampled via locations in which agents meet: schools, workplaces, households and leisure-time. After being infected, agents go through a detailed disease and/or patient pathway that depicts the different states of the disease and the treatment of the patient.

The model input consists of a time-line of modelled NPIs; parameters are calibrated using a modified bisection method. For generation of the weekly forecasts, the model is fitted to the 7 day incidence of the new confirmed cases of the last 21 days including a nowcasting correction for the last week to supplement for subsequent registrations.

Results are gathered via Monte-Carlo simulations as the point-wise sample mean of multiple simulation runs. Due to the large number of agents in the model, 8 simulation runs are used which are sufficient to have the sample mean approximate the real unknown mean with an error of less than 1% with 95% confidence (estimated by the Gaussian stopping introduced in [25]).

The model considers uncertainty with respect to the stochastic perturbations in the model by tracing the standard deviation of the Monte Carlo simulations. Parameter uncertainty is considered in form of manually defined best and worst case scenarios. Parameter values of the model are continuously improved and available online [26].

### D. Epidemiological Clockwork Model

The third model is a simplified state space model that traces individuals through the stages of being infected, the latency period, infectious, reported, contained and immunized/deceased; see also SI Appendix B. Based on the ratio between infected and infectious, the true infection rate is calculated and extrapolated in the future using Kalman Filtering.

The model distinguishes between detected an undetected cases and accounts for the import of infected cases (who did not acquire the infection from the calculated number of infectious individuals).

The underlying detection rate, immigration, and the effectiveness of contact tracing are time-varying parameters that also reflect qualitative information such as spikes in the number of reported new infected cases that can be explained by mass test screenings.

Uncertainty can be modelled by varying underlying model parameters and also results from parameter uncertainty in the infection rate extrapolation.

### E. Model harmonization

In order to harmonize the models and generate a single consolidated forecast for the number of new and accumulated positive COVID-19 tests, each model was set up to generate its output in a common data format for each of the nine federal states of Austria. Our forecasts consisted of time series of confirmed cases for each day starting with the number of positive tests at the day of the forecast consortium meeting at 11:59 pm and and ending between 8 and 14 days in the future (over time, we slightly increased the forecast horizon).

Three averaging procedures were considered to generate the joined forecast from the three individual forecasts. These included (1) the point-wise arithmetic mean and two dynamic weighting procedures wherein the timeseries for each model contributes with a (2) continuous or (3) discrete weighting function with values proportional to the accuracy of its most recent forecasts; see SI Appendix C.

Confidence intervals (CIs) for the harmonized model are derived from the empirical forecast error. Until end of September, CIs where derived from the SIR-X model before the method was refined using the empirical forecast error of the harmonized model. More specific, we retrospectively evaluate the ratio of the consolidated forecast and the actual total number of new cases for each day after the prognosis has been made. The distribution of these ratios reasonably approximates a log-normal distribution for each day. The upper and lower limits of the CI are derived from the corresponding percentiles of the empirical distribution of this forecast error.

### F. Hospital bed usage model

Hospital occupancy is modeled in a stock-flow approach. Inflow (admission to ICU and normal wards) is calculated as a ratio of the time-delayed number of reported or projected new infected. Length of stay is modelled to correspond to the observed distribution and determines outflow. Admission rates are scaled in order to fit the current occupancy in all federal states. The scaling parameter (one for each federal state) can thus be interpreted as an effective hospitalisation rate.

Confidence intervals for the occupancy forecast are calculated from the empirical distribution of the forecast error by initial value (as low occupancy figures are, to a larger degree, subject to fluctuation). A technical description is given in the SI, Appendix D.

Model parameters were initially extracted from literature [27] and subsequently calibrated to actual data to better fit the observed time series. A subsequent analysis based on inpatient data from March 2020 to February 2021 revealed that the calibrated model parameters correspond with observed average length of stay. We refer to the supporting information for a full list of model parameters.

## III. RESULTS

### A. Forecasting positive test numbers

We show the results for our rolling forecasts compared with the actual case numbers in Figure 1. For the time period from April 4 to beginning of February 2021, we performed and harmonized weekly forecasts that are visibly as bundles of lines in Figure 1. The first published forecasts coincided closely with the peak of the first epidemic wave in Austria. This can be seen by a gradual flattening of the curve of cumulative case numbers over April. While the models showed a clearly discernible divergence for the first prognosis, the agreement increased over time. The starting points for the early weekly forecasts occasionally lie below the actual cases due to a substantial amount of very late reporting of cases in these early periods of the epidemic. From May until July, the curve showed a linear growth pattern.

**FIG. 1.**
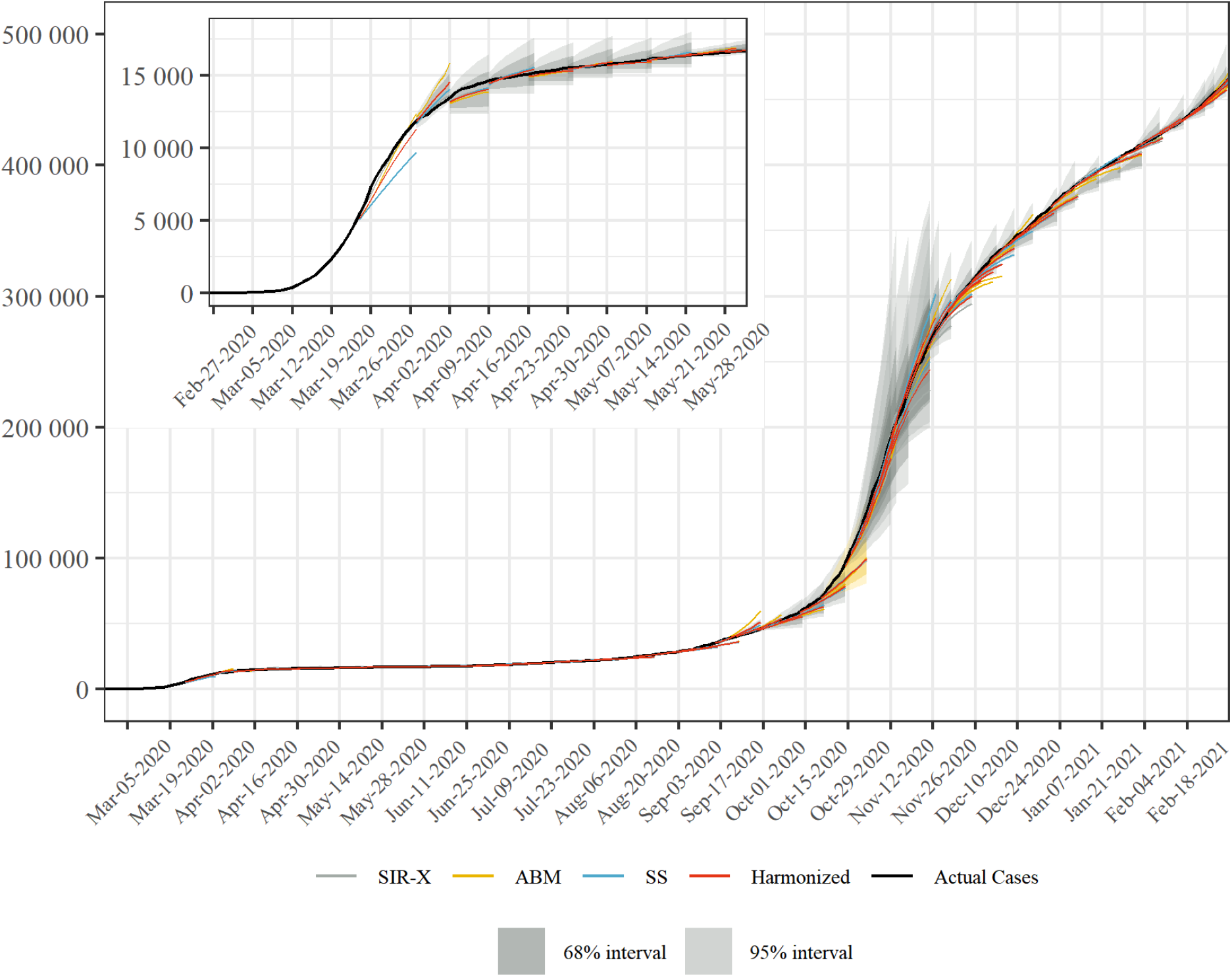
Rolling combined and consolidated out-of-sample forecasts for the number of confirmed cases in Austria. We show the weekly predictions from the three different models, their arithmetic average with it corresponding CI, and the actual case numbers. In the first two weeks, no CI was given. Inset: Individual and harmonized forecasts compared to the actual case number of confirmed cases for the first wave, March to May 2020.

Starting from late August to early September, the onset of the second wave is visible. A critical point was the prognosis for the week until October 25, highlighted in yellow in Figure 1. There, the consolidated forecast clearly underestimated the actual rise in confirmed cases. The prognoses made in the weeks thereafter were more accurate, however, the CI strongly increased due to the less accurate forecasts from the first half of October. A gradual flattening of the curve until end of January can be observed after the steep increase in November, during which the forecasts had a tendency to overestimate the degree of the flattening.

In Figure 2 we compare the model-specific forecast error with the forecast error of the consolidated model, the incidence and the effective reproduction number in Austria. In April 2020, agreement amongst the three models is typically stronger than the agreement with the data, meaning if one model over- or underestimated the actual trend, so did the other models. After the summer, agreement between the three models was larger than in the early phases of the pandemic. Comparing the upper and lower part of this image also shows that none of the models anticipated a spontaneous climb in *R*_*eff*_ in combination with a large number of daily cases in August/September and October. This was particularly distinct and significant at end of October (see the dotted lines in Figure 2).

**FIG. 2.**
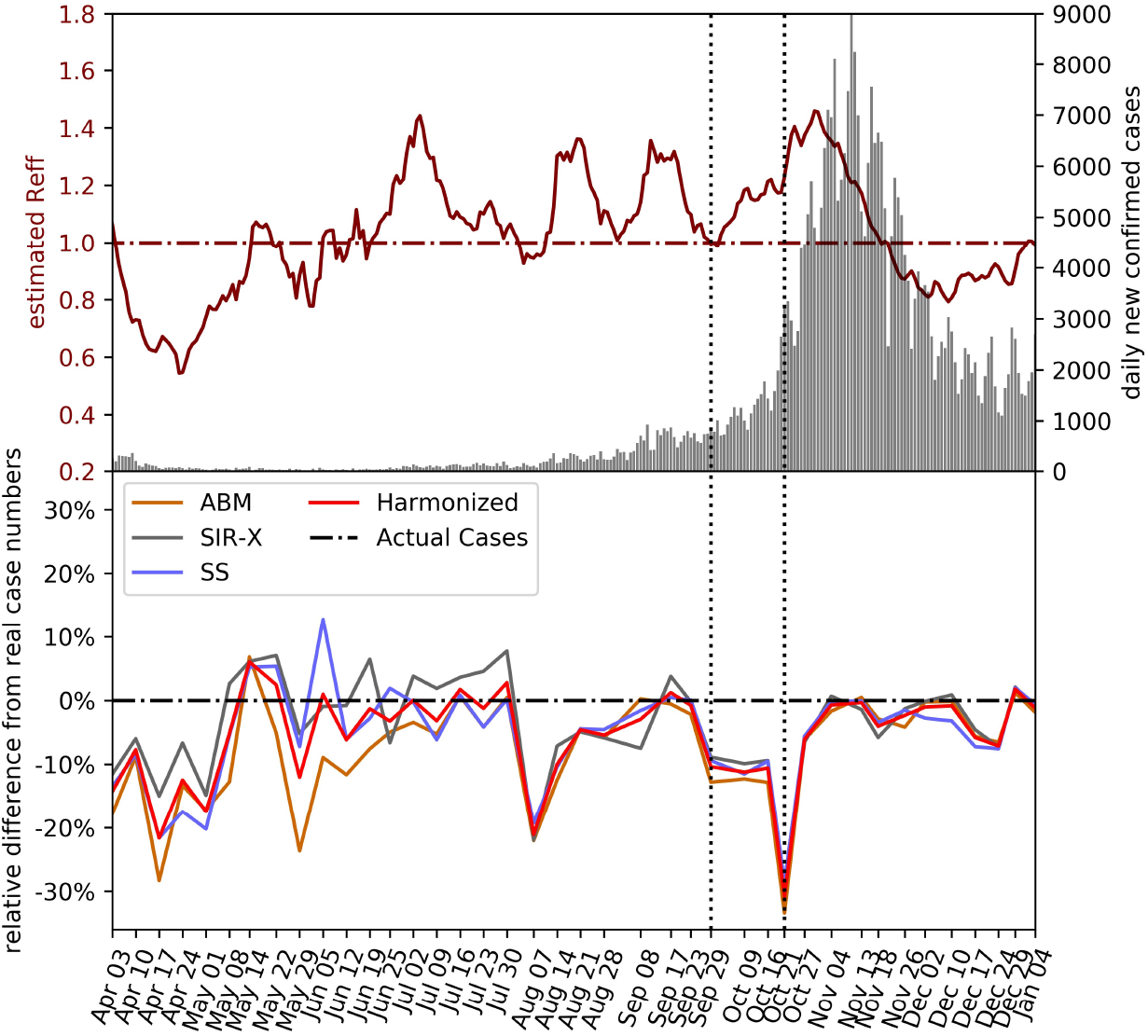
Upper part: daily new confirmed cases and estimated *R*_*eff*_. Lower part: forecasting performance (relative difference) of the individual models and the harmonized mean. Weeks in which the forecasts substantially underestimated the actual case numbers (vertical dotted lines) tend to coincide with high case numbers combined with steep increases in *R*_*eff*_.

We investigated the performance of different averaging procedures that weigh models according to their past performance in terms of their relative difference and error, see Methods. The results are summarized in Table I. Performance weighting procedures yielded only a marginal improvement over simple averaging in terms of forecast accuracy. This further corroborates that agreement amongst the model forecasts is typically higher than their agreement with the observed case numbers.

**TABLE I.** Forecast accuracy with different model harmonization strategies. We consider a naive arithmetic mean (strategy “naive”) as well as a “continuous” and “discrete” function that weighs the models according to their recent performance. The table displays weights and (median) relative errors for three selected weeks and a summary of the forecasts for weeks 2–40. Performance-weighted model harmonization strategies only marginally improved the accuracy of the forecasts compared to a naive averaging.

| Week 10 |  |  |  |  |
| --- | --- | --- | --- | --- |
| strategy | weights for mean |  |  | relative error |
|  | SIR-X | AB | SS |  |
| naive | 0.33 | 0.33 | 0.33 | 10.3% |
| continuous | 0.83 | 0.06 | 0.11 | 7.1% |
| discrete | 0.50 | 0.17 | 0.33 | 9.8% |
| Week 20 |  |  |  |  |
| strategy | weights for mean |  |  | relative error |
|  | SIR-X | AB | SS |  |
| naive | 0.33 | 0.33 | 0.33 | 7.7% |
| continuous | 0.31 | 0.35 | 0.34 | 7.6% |
| discrete | 0.17 | 0.50 | 0.33 | 7.7% |
| Week 30 |  |  |  |  |
| strategy | weights for mean |  |  | relative error |
|  | SIR-X | AB | SS |  |
| naive | 0.33 | 0.33 | 0.33 | 1.2% |
| continuous | 0.40 | 0.16 | 0.44 | 0.6% |
| discrete | 0.33 | 0.17 | 0.50 | 0.4% |
| Weeks 2-40 |  |  |  |  |
| strategy | median relative error |  | IQR |  |
| naive | 7.04% |  | 12.61% |  |
| continuous | 6.73% |  | 12.03% |  |
| discrete | 6.46% |  | 12.08% |  |

### B. Forecasting bed usage

In Figure 3 we show our rolling forecasts for the number of intensive beds in use for COVID-19 patients. There are two peaks in ICU bed occupancy corresponding and delayed with respect to the peaks in case numbers. The red dashed line gives the threshold of 33% of all ICU beds being in use for COVID-19 patients. We highlight again (yellow in Figure 3) the forecast for the week until October 25, in which we severely underestimated the developments. After adjusting the models for these developments, the next forecast projected that the 33% threshold would likely be crossed in two to three weeks. After the second peak passed, the model showed a tendency to overestimate the pace at which patients would be released from ICU.

**FIG. 3.**
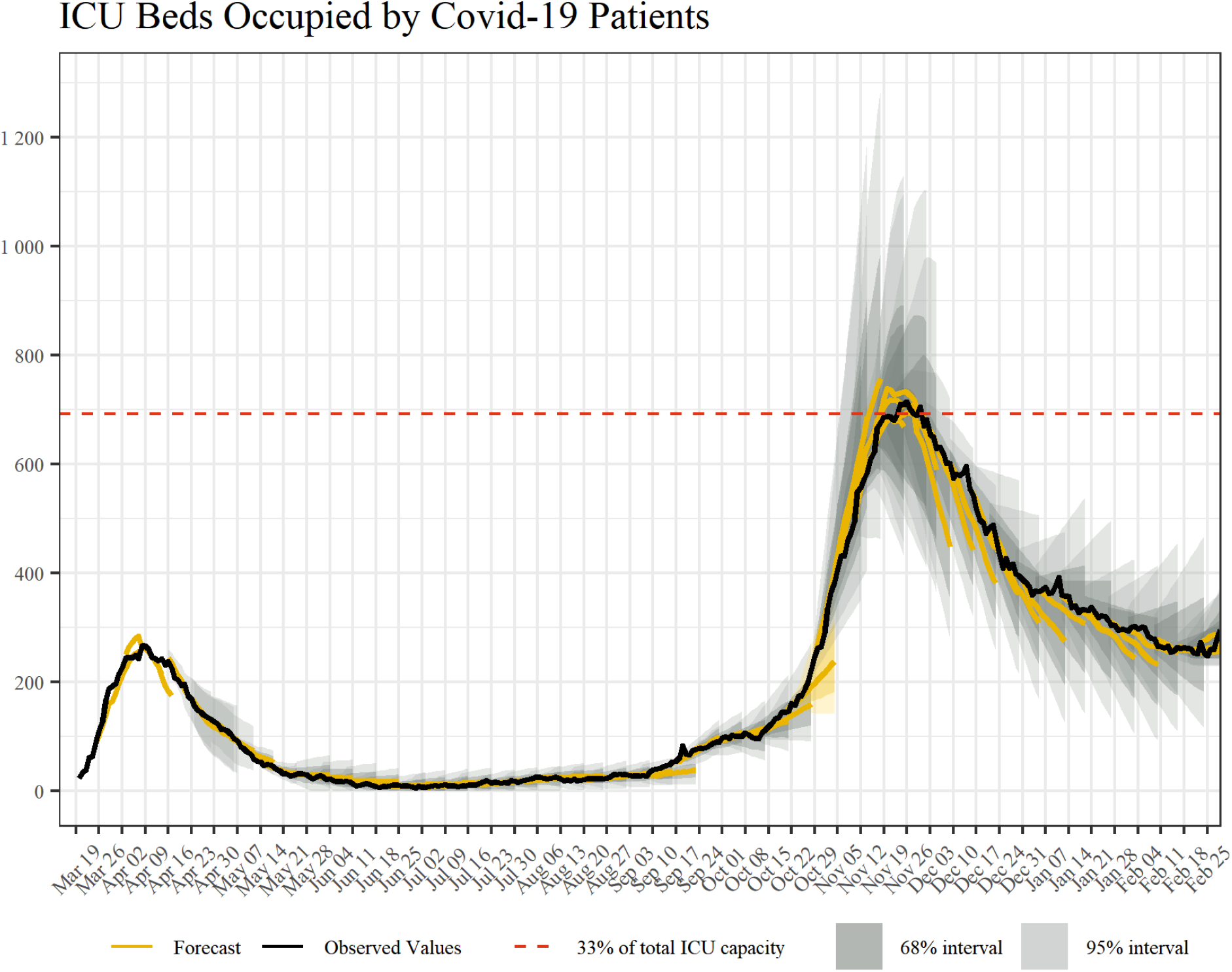
Rolling combined and consolidated out-of-sample forecasts for the number of intensive care beds currently in use for COVID-19 patients including corresponding CI, the actual numbers of beds occupied, and 33% of total ICU capacity as a reference.

### C. Reporting of the forecasts

We developed a standard reporting template used to communicate our forecasts to other stakeholders and decision-makers, see Figure 4. These visual reports showed our forecasts for cases and hospital occupancy, as well as information on the effective reproduction number. The visual reports are complemented by a brief synopsis of the researcher’s appraisal of the current situation and particularities of the most recent forecast. We highlight what drives our results and illustrate the nature of the underlying uncertainty, such as the role of tourists returning from risk areas in late August. Furthermore, the researchers are at disposal for any questions that members of the health ministers’ office or the regional crisis management units may have.

**FIG. 4.**
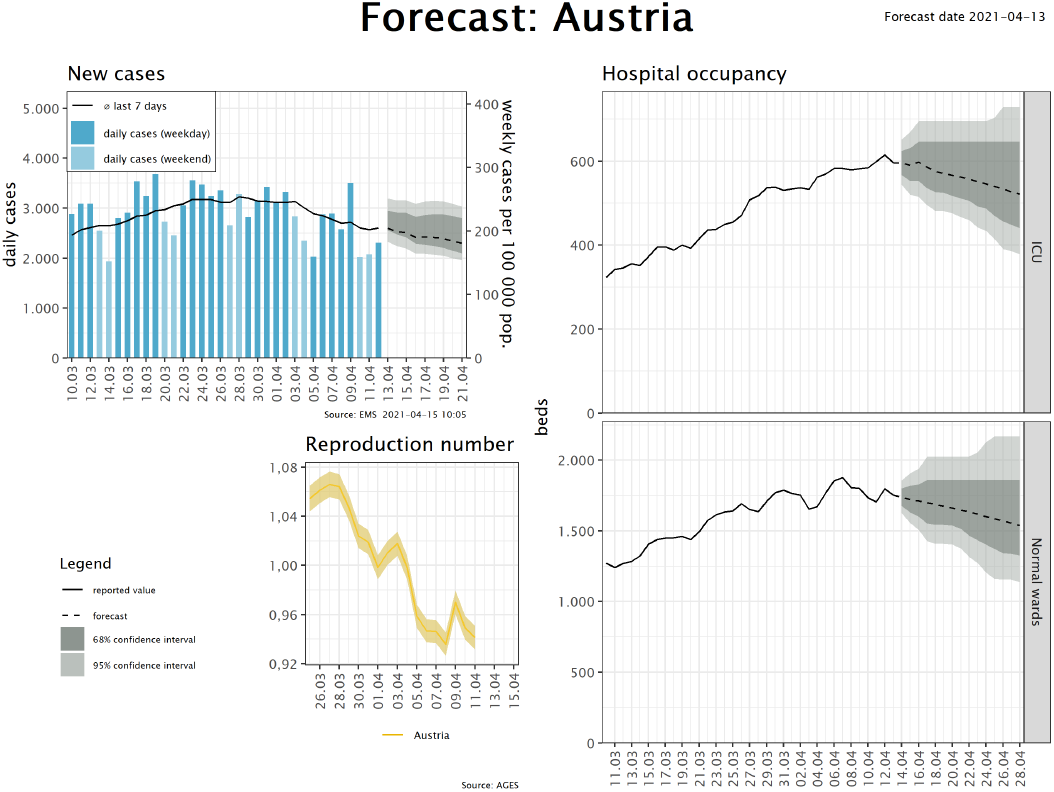
Example for a reporting template of our out-of-sample forecasts. The visual reports consist of four panels. First, we report the daily number of cases in absolute terms and the weekly case numbers per 100 000 population as well as the forecasts for the weekly case numbers, including CI. Additional panels show the the effective reproduction number and the forecasts for intensive and normal care beds occupancy with COVID-19 patients.

The first panel in Figure 4 provides an outlook for the expected developments in weekly cases (per 100 000 population). Due to known weekday effects, we do not give forecasts for daily case numbers. Expected hospital occupancy is given in the second panel. All forecasts are supplemented with 68% and 95% confidence intervals. The forecasts are also presented in the weekly sessions of the Austrian Corona Commission, an advisory body to the minister of health tasked with assessing the epidemiological risk in Austria. The expected occupancy rates of ICU is an indicator in assessing the risk of health system overburdening and thus directly contributes to the classification of Austrian regions according to this risk, which in turn informs recommendations on easing or strengthening control measures [6].

## IV. DISCUSSION

Considering the impact of COVID-19 policy measures on economic and social life, any related decision support needs to be done with caution. Our approach considers the high impact of COVID-19 forecasts by (1) focusing on monitoring rather than long-term prognosis and (2) consolidation of three different “model opinions” which not only improved the quality of the short-term forecast, but also distributed the responsibility of the decision support on multiple teams.

From the very beginning of our work as consortium, we decided to publish only short-term forecasts and to refrain from publishing long-term scenarios. Due to the multiplicative growth of uncertainties in epidemiological models, accurate forecasts are typically only possible over a time horizon of several days [28–31]. There is no meaningful way to estimate the uncertainty for long term scenarios that span over several weeks, months or even years. For policy-makers and non-technical experts, the distinction between a prognosis with a defined level of certainty and a hypothetical what-if experiment is hard to communicate.

Our forecasts provided evidence for the expected total number of daily infections and hospitalised cases including appraisals of uncertainty via forecast intervals. This is in contrast to what has come to be known as “worst case coronavirus science”, i.e. the communication of worst case scenarios as baselines in the public pandemic management strategy. For instance, the UK policy change toward adopting more drastic NPIs on March 23 was based on worst case scenarios created by the Imperial College COVID-19 Response Team that within the current policy regime 250,000 deaths were to be expected. In a press conference in April, the Austrian chancellor publicly stated that soon “everyone will know someone who died because of COVID-19”, based on an external SIR-model-based worst case scenario that contained a death toll of 100,000 people (1.1% of the Austrian population) [32].

Such scenarios are problematic due to the high levels of uncertainty of long-term (multiple weeks and months) case number forecasts; a generic feature of mathematical epidemiological models which has been put under the spotlight by the COVID-19 outbreak [29, 31, 33, 34]. Based on our results, we argue that a main benefit of epidemiological models comes from their use as short-term monitoring systems. The models are typically calibrated to the infection dynamics of the last couple of days or weeks and forward project this dynamic based on epidemiological parameters often assumed to be fixed. If a short-term forecast is accurate this means that infection numbers have continued as expected, based on the recent trend and accurate assumptions. In our forecasts, as shown in Figure 2, this holds true for time periods from May to July 2020 and from November 2020 onwords. If the short term forecast severely over- or underestimates the observed dynamics, one should inquire more closely what might have caused this change.

Inaccurate short term forecasts signal a change in the epidemiological situation that needs to be explained. This occurred, for instance, when our forecasts in August consistently underestimated the observed case numbers. At this point, contact tracing data revealed a growing number of travel associated cases mainly from hotspots in South Eastern Europe contributed substantially to this unexpected increase in infections. In response, novel border restrictions for persons entering Austria from these countries were put in place at the end of August. During the summer 2020, infection numbers increased from around 20 confirmed cases per day to about 200 cases, mostly driven by patients aged below 40 years. Consequently, the number of severe COVID-19 cases remained low and the effective rate to require intensive care dropped to one percent and below. The situation changed qualitatively in September, when not only case numbers started to soar again, but also hospital admissions increased much more strongly than projected.

Our analysis revealed that the driver behind the observed above-forecast ICU occupancy was above-forecast total case numbers, while age-specific ICU rates remained constant in our models. In other words, the bed usage forecasts were inaccurate because of the infection number forecast, but not because the characteristics of the detected cases changed in terms of severity (e.g., age or more symptomatic or severe cases).

At the onset of the second COVID-19 wave in Austria in October 2020 the forecasts influenced policy debate and contributed to the decision of implementing a second lockdown. The forecast of October 30 predicted an increase in ICU occupancy by COVID-19 patients from 263 to 681 (34% of total ICU capacity of 2,007 beds) within the next 15 days and stressed that capacity limits of around 700 to 800 ICU beds dedicated to COVID-19 patients may be exceeded by mid to end of November if this trend would be unbroken. Additionally, the heterogeneous trend across Austrian federal states -– leading to regions with COVID-19 ICU occupancy rates of more than 50% -– was emphasised [35]. In a speech in the

Main Committee of the National Council the Austrian Federal Minister of Health emphasized the dynamic of the second wave. Referring to the forecast of October 30 which predicted an increase in daily case numbers up to 6,300 by November 7 and a critically high level of ICU occupancy rate, the Minister called for a second lockdown [35, 36]. While actual ICU occupancy remained below the forecast (e.g. 585 vs. 681 on November 14) the call for action turned out to be appropriate as the Austrian intensive care system operated around its capacity limit with a maximum of 709 ICU beds occupied by COVID-19 patients on November 25 according to available information.

After relaxing NPIs during December 2020 the forecasts results also contributed to the third lock-down after Christmas where persistently high levels of ICU occupancy rates were predicted and concerns regarding the seasonal increase in contacts which may lead to a further increase in occupancy rates were raised [37].

One might question whether complex epidemiological models are indeed necessary for such a short-term forecast-based monitoring system or whether the use of simpler models could not serve a similar purpose. Indeed, models that are not of the compartmental SIR type but use other prediction algorithms, ranging from ARIMA [38, 39] to deep learning [40, 41], have also been applied to forecast the SARS-CoV-2 pandemic. The advantage of using compartmental epidemiological models is that they also provide a mechanistic description of why changes in the current epidemiological situation are occurring. In particular, as a consortium we were frequently asked by policy-makers to provide estimates for possible future epidemiological developments given a certain NPI would be implemented in a few days or not. To answer such requests, it is beneficial to use models allowing to directly simulate the effects of hypothetical interventions. Such questions can be even more reliably and consistently answered if the same model is used to produce short-term forecasts as a baseline scenario and a hypothetical scenario assuming the implementation of a new NPI.

Our forecast-based decision support comes with limitations. First, the weekly prognosis is partly based on shared data from the Ministry of Health and the Ministry of Internal Affairs which comes with quality and reporting bias limitations. Moreover, even though the consortium has access to the most accurate and up-to-date data about the epidemic in Austria, a lot of information required for valid epidemiological forecasting is not available or only available with considerable delay, since adequate reporting systems are not in place; e.g., the fraction of undetected persons due to asymptomatic disease progression. Further, our forecast is based on simulation models which are generally subject to errors that come from abstraction and simplification of the real system. Through the harmonized handling of three models with entirely different approaches we attempted to reduce such structural uncertainties. Finally, our decision support framework is mostly limited by its political and public visibility. According to our experience, our forecast was of special public and policy interest in periods of rapid movements but also had a confirmatory effect in times of decreasing case numbers or slow growth with respect to taken policy measures.

In conclusion, we argue that modellers need to be cautious and responsible in communicating the sometimes strongly speculative nature of their results and their uncertainties to politicians and the public. Short-term epidemiological models can be valuable ingredients of a comprehensive monitoring and reporting system to detect epidemiological change points and thereby inform decisions to ease or strengthen governmental responses.

## Data Availability

Data is available online.

https://www.data.gv.at/covid-19/

## V. ACKNOWLEDGMENTS

We thank Reinhild Strauss, Gabriela El Belazi, Lukas Richter, Daniela Schmid, Erich Neuwirth, and Uwe Siebert for helpful and stimulating discussions.

## VII. AUTHOR CONTRIBUTIONS

MB, CR, and NP developed and operated the Agent-Based SEIR Model. MZ, LR, FB, and HO developed and operated the Epidemiological Clockwork Model. ST and PK developed and operated the Extended SIR-X Model. MB, PK, MZ, LR and FB developed the hospital bed occupancy model. PK wrote the first draft of the article. MB, MZ, LR, FB and CR contributed to the writing. All authors reviewed and edited the manuscript.

## SUPPORTING INFORMATION

## Appendix A: Details on the Extended SIR-X Model

The SIR-X model [17] was originally devised to model the transition from exponential to sub-exponential growth due to the implementation of non-pharmaceutical interventions (NPIs). With the entry into containment 2.0, i.e. the taking back of NPIs, a couple of modifications are therefore needed to the original model. In particular, the process of “quarantining” susceptibles has to be added. In the following we give a description of how we extended the baseline SIR-X model in order to implement such processes. We also discuss additional extensions to the model, such as introducing age-structured populations and multiple calibration periods. Note that this document only discusses aspects of the model implementation not described or treated different than in the original model description.

### 1. Compartments for quarantined infected and susceptibles

Here we give the extended SIR-X model to account for scaling back NPIs. The baseline model includes two different types of NPI. First, there are NPIs that act on the susceptible population (social distancing, home office, etc.). Second, there are NPIs that act on the infected population, in particular an accelerated detection of cases (e.g., testing and contact tracing). Clearly, scaling back of the NPIs affects primarily the first type of NPIs, while it might be reasonable to expect that NPIs targeting the infected population might even increase in effectiveness.

The baseline SIR-X model is of the following form,

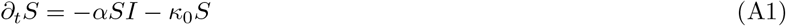

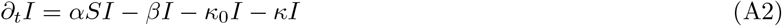

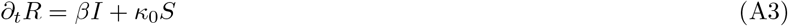

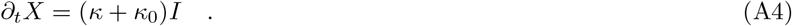

We now introduce two extensions, namely (i) having two compartments of locked down individuals (susceptibles, *X*^*S*^, and infecteds, *X*^*I*^) and (ii) introducing a scaling back of NPIs affecting susceptibles encapsulated in the rate *κ*_1_ ≥ 0. The extended SIR-X model is then of the following form,

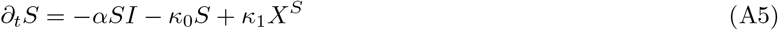

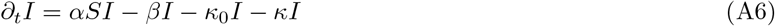

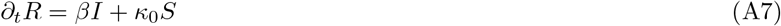

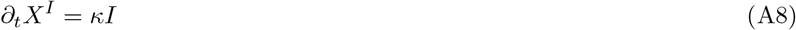

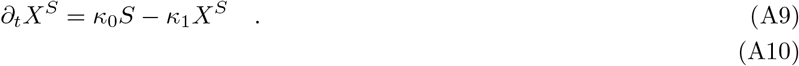

There are two notable differences now. First, the compartment *X*^*I*^ is the *cumulative number of confirmed cases*, it will be used to calibrate the model. It is imperative to note that the model was explicitly designed to make statements concerning *X*^*I*^. A couple of further straight-forward extensions could be introduced to model, for instance, also recovery within the active *X*^*I*^ compartment. Such extensions would further add to the model complexity while providing no value at all for modelling the development of *X*^*I*^. Secondly, the compartment *X*^*S*^ is now an explicit model representation of locked down or socially distanced susceptibles. The parameter *κ*_0_ gives the inflow to this compartment from the susceptibles (strength of corresponding NPIs), *κ*_1_ gives the outflow (how fast people increase their levels of social contacts back to normal).

### 2. Age structure

We include an age structure in the model in the ususal way. All compartments (*S, I, R, X*^*I*^, *X*^*S*^) become vector-valued, so do the rates *κ*_0_, *κ*_1_, *κ*, and *β*. The parameter *α* becomes a matrix *α*_*ij*_ giving the likelihood that a susceptible of age group *j* will be infected by an infected from age group *i*. Entries in *α* have been calibrated using mobile phone data, where we assume that they are proportional to the probability that a call will take place between individuals of age group *i* and *j*. The spectral radius of *α* is chosen in accordance with [17].

### 3. Calibration

As already mentioned, the model is calibrated via the timeseries of the cumulative number of confirmed cases *X*^*I*^ in each federal state of Austria. The basic calibration procedure follows [17], i.e. we solve the model for solutions to the parameters *κ* and *κ*_0_, as well as the initial condition *I*(*t* = 0) via a trust region reflective algorithm (MatLab’s lsqnonlin function). Calibration takes place in different time windows that roughly represent the different phases of the epidemic in Austria. The first phase lasts from *t* = 0 to end of March and encompasses the “first wave”. With beginning of April, Austria moved into a containment phase characterized by less than hundred new confirmed cases per day. The second calibration phase ends mid-June where the daily cases started to increase again with most days showing more than hundred new cases. The third calibration phase lasts until the end August, when infection numbers started to increase again, after which the fourth calibration phase commenced.

## Appendix B: Details on the Epidemiological Clockwork Model

The epidemiological stage model tracks individuals through the stages “infected”, “infectious”, “reported”, “isolated”, and “immunized”. It aims to isolate the true infection rate via augmenting reported case numbers with time-constant epidemiological parameters and time-varying assumptions on detection rate, isolation rate, and number of imported cases. This infection rate is then extrapolated using exponential smoothing models in order to forecast future case numbers.

### 1. Data preparation

In a first step, assumptions on detection and isolation rates are made based on the cluster analysis. For example, if a large share of new reported cases is attributed to a single workplace setting where the whole staff was tested, we assume that contact isolation and detection rate are high at the day these cases were reported, and were lower in days before the mass testing. We furthermore subtract sporadically imported cases as reported in the cluster analysis. This serves to isolate domestically acquired infections for the calculation of the infection rate.

In a second step, case numbers are cleared in a nowcasting procedure, where expected delays and weekend effects are accounted for based on historic values of the relevant federal states. We use exponential smoothing to identify seasonality, error and level of the case numbers for each federal state.

In a third step, assumptions on the risk of importing infectious cases are made, i.e. people who have not acquired the infection domestically but increase the number of infectious.. This figure is parameterised to account for size of federal states and adjusted to reflect travel patterns in risk areas. For example, this value was increased to account for a spike in reported cases that were traced back to travellers from Croatia.

### 2. Model parameters

Fixed parameters

- Duration infected before infectious: *d*_1_ = 1 day
- Duration infectious before reported: *d*_2_ = 1 day Duration infectious for non-severe cases: *d*_3_ = 6 days Latency period is therefore 2 days. This value was initially set to 3 days but was reduced owing to evidence that average latency period is shorter. Note that transmission relies on both pre-symptomatic detected cases as well as non-detected cases, who may or may not have mild symptoms.

Variable parameters (default values)

- Background infection risk_*t*_ = population*/*100, 000
- Detection rate: new reported cases*t/*new total cases_*t*_ = *r*_1_ = 1*/*5
- Effectiveness of contact isolation: new isolated cases_*t*_*/*new reported cases*t* = *r*_2_ = 2*/*5

Those given values are standard parameterisations that are adjusted to account for specific circumstances. In the standard case, there would be four additional undetected cases for every reported case, and on average two infected people would be quarantined for every reported case.

### 3. Mathematical model

The model calculates the number of individuals in each stage according to the following difference equations,

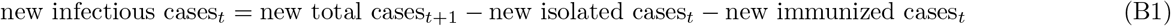

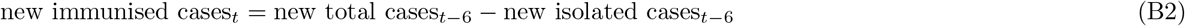

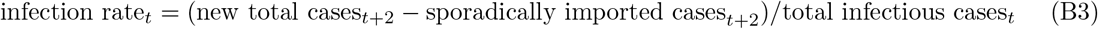

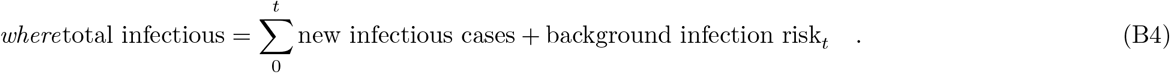

### 4. Trend extrapolation

In a last step, the isolated true infection rate is forecasted in an exponential smoothing model (R, package ‘forecast’). Owing to the tendency of the disease to progress linear instead of exponential [12], all trends are damped.

### 5. Strengths and Limitations

In the Epidemiological Stage Model, isolation of the true infection rate is by a large degree driven by researcher assumptions on the key time-varying variables of imported cases and detection rates. This follows the understanding that reported case numbers should be assessed with available additional information before being further processed by mathematical models. Known sources of error, such as non-detection of cases, travel activity of infected or non-randomised testing will affect, unless these sources of bias are time-constant and the relevant sample size is large. The otherwise simple mechanic of the model facilitates attribution of case numbers to such causes as time of reporting determines day of infection. Of course, researchers may err as well as model when trying to attribute observed patterns to causes, or parameters. The Epidemiologic Clockwork Model will perform better than more data-driven models if known particularities in the time-series of reported cases play a substantial role in the course of transmission. A limitation of the model is that due to the nature of the information used in assessing the time series of reported cases, which is often qualitative in nature, the data clearing process is not transparent. Ongoing improvements of the model will address these shortcomings as the pandemic progresses.

## Appendix C: Combining the three models into a consolidated forecast

In summary, three strategies have been evaluated in terms of their forecast error. Let 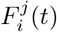 denote the forecasts for the total number of COVID-19 cases on day *t* for model *j* for runs made in week *i*,

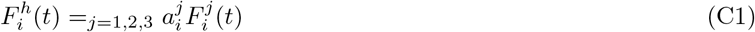

the harmonized forecast, and *R*_*i*_ the corresponding reported number, then the following three strategies have been investigated to calculate the weights 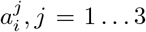. Note that, here and in the following, *j* is an index and not an exponent.

- **Naive average**. This strategy describes a static arithmetic average of the forecasts.

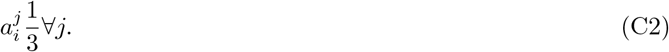
- **Continuously weighted dynamic mean**. This strategy describes a dynamically weighted average. The weights are determined from the forecasting errors of the previous three weeks.

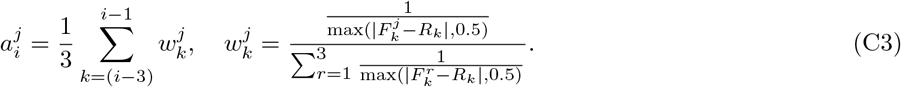
- **Discrete weighted dynamic mean**. In contrast to the continuous weighting, the weights are determined by a step function as follows,

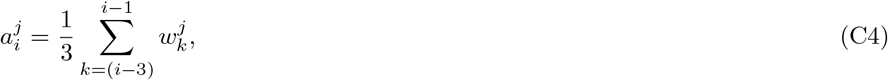

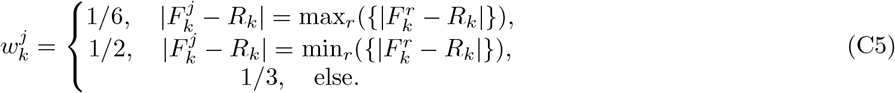

## Appendix D: Confidence Intervals

Validation of the model is now conducted by evaluating the model’s forecast error in each federal state over the last forty days. We compute the empirical distribution of the daily ratios beteen the fitted and observed numbers of cumulative confirmed cases. This gives us a 68% and 95% confidence interval (CI) for the expected accuracy of the model for the prediction for the next day. Following a similar strategy as reported in the online implementation of the baseline model [42], we than recalibrate the model in the last (third) calibration phase by doing as if the upper and lower bounds of these CIs are the actual data points for the next day. The CIs for the forecast are obtained from the forecasts that start from these “virtual” observations.

For the confidence intervals of the ICU and hospital occupancy we decided to apply a different strategy because the fluctuations of the occupancy numbers played a much higher role for the error than the parameter uncertainty. In specific, an almost linear relation between the level of uncertainty and the squareroot of the occupancy on the first prognosis day could be observed.

We regard the tuples 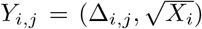. Hereby, Δ_*i,j*_ denotes the logarithmic error 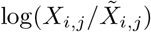 between the reported occupancy *X*_*i,j*_ and the forecast occupancy 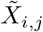 of day *j* in prognosis week *i*. Furthermore 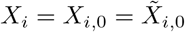 denotes the occupancy on the day of the prognosis in week *i*.

Furthermore, the Tuples *Y*_*i,j*_ are evaluated in by a 2-dimensional kernel-density estimation (KDE). Gaussian kernels and a standard Scott’s Rule [43] are used for the KDE. A kernel function 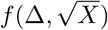 results.

To generate confidence intervals, the marginal distributions

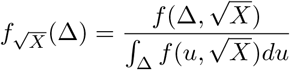

are calculated numercially. Finally, the numerically calculated percentiles of 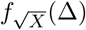 result in the required confidence bands for the logarithmic error, given a certain occupancy *X* on the day of the prognosis.

## References

[1] C. L. Correa-Martínez, S. Kampmeier, P. Kümpers, V. Schwierzeck, M. Hennies, W. Hafezi, J. Kühn, H. Pavenstädt, S. Ludwig, and A. Mellmann, “A pandemic in times of global tourism: superspreading and exportation of covid-19 cases from a ski area in austria,” Journal of clinical microbiology, vol. 58, no. 6, 2020.

[2] P. Kreidl, D. Schmid, S. Maritschnik, L. Richter, W. Borena, J.-W. Genger, A. Popa, T. Penz, C. Bock, A. Bergthaler, et al., “Emergence of coronavirus disease 2019 (covid-19) in austria,” Wiener klinische Wochenschrift, pp. 1–8, 2020.

[3] A. Remuzzi and G. Remuzzi, “Covid-19 and italy: what next?,” The Lancet, 2020.

[4] A. De Giorgio, “Covid-19 is not just a flu. learn from italy and act now,” Travel Medicine and Infectious Disease, 2020.

[5] “Änderung des epidemiegesetzes 1950, des tuberku-losegesetzes und des covid-19-maßnahmengesetzes,” 2020.

[6] GÖG/AGES, “Manual corona-kommission,” 2021.

[7] A. Desvars-Larrive, E. Dervic, N. Haug, T. Niederkro-tenthaler, J. Chen, A. Di Natale, J. Lasser, D. S. Gliga, Roux, A. Chakraborty, et al., “A structured open dataset of government interventions in response to covid-19,” Scientific Data, vol. 7, no. 285, 2020.

[8] “Amtliches dashboard covid19.” https://info.gesundheitsministerium.at/. Accessed: 2020-09-09.

[9] “Covid-19 information page by ages.” https://www.ages.at/en/wissen-aktuell/publikationen/epidemiologische-parameter-des-covid19-ausbruchs-oesterreich-20202021/. Accessed: 2020-08-18.

[10] S. Thurner, R. Hanel, and P. Klimek, Introduction to the theory of complex systems. Oxford University Press, 2018.

[11] D. R. L. Sardo, S. Thurner, J. Sorger, G. Duftschmid, G. Endel, and P. Klimek, “Quantification of the resilience of primary care networks by stress testing the health care system,” Proceedings of the National Academy of Sciences, vol. 116, no. 48, pp. 23930–23935, 2019.

[12] S. Thurner, P. Klimek, and R. Hanel, “A network-based explanation of why most covid-19 infection curves are linear,” Proceedings of the National Academy of Sciences, 2020.

[13] M. R. Bicher, C. Rippinger, C. Urach, D. Brunmeir, U. Siebert, and N. Popper, “Agent-based simulation for evaluation of contact-tracing policies against the spread of sars-cov-2,” medRxiv, 2020.

[14] N. Popper, F. Endel, R. Mayer, M. Bicher, and B. Glock, “Planning Future Health: Developing Big Data and System Modelling Pipelines for Health System Research,” SNE Simulation Notes Europe, vol. 27, pp. 203–208, Dec. 2017.

[15] M. Bicher, C. Urach, and N. Popper, “GEPOC ABM: A Generic Agent-Based Population Model for Austria,” in Proceedings of the 2018 Winter Simulation Conference, (Gothenburg, Sweden), pp. 2656–2667, IEEE, 2018.

[16] C. Rippinger, M. Bicher, C. Urach, D. Brunmeir, N. Weibrecht, G. Zauner, G. Sroczynski, B. Jahn, N. Mühlberger, U. Siebert, and N. Popper, “Evaluation of undetected cases during the COVID-19 epidemic in Austria,” BMC Infectious Diseases, vol. 21, p. 70, Jan. 2021.

[17] B. F. Maier and D. Brockmann, “Effective containment explains subexponential growth in recent confirmed covid-19 cases in china,” Science, vol. 368, no. 6492, pp. 742–746, 2020.

[18] M. Gatto, E. Bertuzzo, L. Mari, S. Miccoli, L. Carraro, R. Casagrandi, and A. Rinaldo, “Spread and dynamics of the covid-19 epidemic in italy: Effects of emergency containment measures,” Proceedings of the National Academy of Sciences, vol. 117, no. 19, pp. 10484– 10491, 2020.

[19] G. Giordano, F. Blanchini, R. Bruno, P. Colaneri, A. Di Filippo, A. Di Matteo, and M. Colaneri, “Modelling the covid-19 epidemic and implementation of population-wide interventions in italy,” Nature Medicine, pp. 1–6, 2020.

[20] S. Flaxman, S. Mishra, A. Gandy, H. J. T. Unwin, T. A. Mellan, H. Coupland, C. Whittaker, H. Zhu, T. Berah, J. W. Eaton, et al., “Estimating the effects of non-pharmaceutical interventions on covid-19 in europe,” Nature, vol. 584, no. 7820, pp. 257–261, 2020.

[21] A. Arenas, W. Cota, J. Gomez-Gardenes, S. Gómez, C. Granell, J. T. Matamalas, D. Soriano-Panos, and B. Steinegger, “A mathematical model for the spatiotemporal epidemic spreading of covid19,” MedRxiv, 2020.

[22] E. Estrada, “Covid-19 and sars-cov-2. modeling the present, looking at the future,” Physics Reports, 2020.

[23] https://www.sozialministerium.at/Informationen-zum-Coronavirus/Neuartiges-Coronavirus-(2019-nCov)/COVID-Prognose-Konsortium.html.

[24] G. Heiler, T. Reisch, J. Hurt, M. Forghani, A. Omani, A. Hanbury, and F. Karimipour, “Country-wide mobility changes observed using mobile phone data during covid-19 pandemic,” arXiv preprint arXiv:2008.10064, 2020.

[25] M. Bicher, M. Wastian, D. Brunmeir, M. Rößler, and N. Popper, “Review on Monte Carlo Simulation Stopping Rules: How Many Samples Are Really Enough?,” in Proceedings of the 10th EUROSIM Congress on Modelling and Simulation, (Logrono, Spain), In Print, July 2019.

[26] http://www.dwh.at/en/projects/covid-19/.

[27] https://www.rki.de/DE/Content/InfAZ/N/Neuartiges_Coronavirus/Steckbrief.html.

[28] M. Castro, S. Ares, J. A. Cuesta, and S. Manrubia, “The turning point and end of an expanding epidemic cannot be precisely forecast,” Proceedings of the National Academy of Sciences, 2020.

[29] W. C. Roda, M. B. Varughese, D. Han, and M. Y. Li, “Why is it difficult to accurately predict the covid-19 epidemic?,” Infectious Disease Modelling, 2020.

[30] N. P. Jewell, J. A. Lewnard, and B. L. Jewell, “Predictive mathematical models of the covid-19 pandemic: Underlying principles and value of projections,” Jama, vol. 323, no. 19, pp. 1893–1894, 2020.

[31] I. Holmdahl and C. Buckee, “Wrong but useful—what covid-19 epidemiologic models can and cannot tell us,” New England Journal of Medicine, 2020.

[32] https://www.oesterreich.gv.at/dam/jcr:a9ba0dbb-fc05-4b6f-a7cb-ecb8b6842364/Executive%20Summary%20Covid19%20v2.pdf.

[33] J. P. Ioannidis, S. Cripps, and M. A. Tanner, “Forecasting for covid-19 has failed,” International journal of forecasting, 2020.

[34] N. N. Taleb, Y. Bar-Yam, and P. Cirillo, “On single point forecasts for fat-tailed variables,” International Journal of Forecasting, 2020.

[35] “Sachverhalt und begründungen zur 2. novelle der covid-19-schutzmaßnahmenverordnung.” https://www.sozialministerium.at/dam/jcr:dfbea104-f4a8-40ab-8a2c-bf7a1cd41458/20201112_Sachverhalt%20und%20Begr%C3%BCndungen%20zur%202.%20Novelle%20der%20Covid-19-%20Schutzma%C3%9Fnahmenverordnung.pdf. Accessed: 2021-01-15.

[36] “Hauptausschuss genehmigt covid-19-schutzmaßnahmenverordnung.” https://www.parlament.gv.at/PAKT/PR/JAHR_2020/PK1114/index.shtml. Accessed: 2021-01-15.

[37] “Hauptausschuss genehmigt covid-19-notmaßnahmenverordnung.” https://www.parlament.gv.at/PAKT/PR/JAHR_2020/PK1468/index.shtml. Accessed: 2021-01-15.

[38] G. Perone, “An arima model to forecast the spread of covid-2019 epidemic in italy,” arXiv preprint arXiv:2004.00382, 2020.

[39] D. Benvenuto, M. Giovanetti, L. Vassallo, S. Angeletti, and M. Ciccozzi, “Application of the arima model on the covid-2019 epidemic dataset,” Data in brief, vol. 29, p. 105340, 2020.

[40] F. Shahid, A. Zameer, and M. Muneeb, “Predictions for covid-19 with deep learning models of lstm, gru and bilstm,” Chaos, Solitons & Fractals, vol. 140, p. 110212, 2020.

[41] A. Zeroual, F. Harrou, A. Dairi, and Y. Sun, “Deep learning methods for forecasting covid-19 time-series data: A comparative study,” Chaos, Solitons & Fractals, vol. 140, p. 110121, 2020.

[42] “Sir-x model, event horizon.” http://rocs.hu-berlin.de/corona/docs/forecast/model/. Accessed: 2020-08-18.

[43] D. W. Scott, Multivariate density estimation: theory, practice, and visualization. John Wiley & Sons, 2015.

